# Better Sleep, Better Minds: Sleep as a Key to Cognitive Performance and Quality of Life in Aging

**DOI:** 10.1101/2025.05.16.25327786

**Authors:** Marta Aliño, Francisco Molins, Mireia Molins, Lorena González, Aranzazu Duque, Patricia Mesa-Gresa

## Abstract

Cognitive decline can significantly affect the quality of life of healthy older adults. There is an age-associated cognitive decline, but there is also evidence that sleep quality is also affected by the aging process. The primary aim of this study is to evaluate whether sleep quality moderated the relationship between cognitive performance and quality of life on a sample of healthy older adults, as well as to evaluate differences between men and women in cognitive performance, sleep quality and perceived quality of life. A final sample of 50 participants was included with a mean age of 66.57 years (71.43% women). Participants completed a battery of questionnaires, including the Fototest (cognitive performance), the Pittsburgh Sleep Quality Index (PSQI) (sleep quality), and European Quality of Life 5 Dimensions (EuroQoL-5D) (perceived quality of life). A correlation analysis was performed for Fototest, EuroQoL-5D, and PSQI scores and the three variables were significantly related in the global sample. Women maintained a similar pattern of relationships between the main variables, whereas none of the correlations reached statistical significance for men. Also, a multiple linear regression analysis was performed and both, cognitive performance and sleep quality, significantly predicted perceived quality of life. Furthermore, a moderation model analysis was conducted explaining approximately 42.2% of the variance in quality of life (R^2^ = 0.42), *F*(4, 37) = 6.76, *p* < .001. Specifically, results showed that sleep quality not only directly influences perceived quality of life but also moderates the relationship between cognitive performance and quality of life. In conclusion, it would be interesting to promote further research on the weight that sleep quality may have when implementing a cognitive intervention program in a sample of older adults, as well as to have a forecast of the impact that such intervention will have on the sample, considering the quality of sleep of the individuals.

## Introduction

Intact cognitive function is crucial to the standard of living for individuals progressing from middle to late adulthood. Age-related cognitive decline is a natural process that affects people as they age. Studies have shown that this decline can begin to manifest itself as early as middle age and affects various cognitive functions, including memory, attention, processing speed and executive functions (Guan et al., 2020; Krivanek et al., 2021). Firstly, this cognitive decline can significantly affect the quality of life of healthy older adults (He et al., 2023). But, in addition, difficulties in remembering information, concentrating on tasks or making decisions can generate frustration, anxiety and affect people’s ability to perform daily activities (Wennberg et al., 2017; Casagrande et al., 2022). This loss of autonomy and independence can lead to social isolation and decreased self-esteem, also negatively impacting quality of life (Brandt et al., 2022; Cristopher and Facal, 2023).

Following different studies, several factors can influence cognitive performance, and thus the quality of life of older adults. These include genetic factors (Rainey-Smith et al., 2018), chronic diseases such as hypertension and diabetes (Rainey-Smith et al., 2014; Meng et al., 2023), educational level (Kaur et al., 2019; Lövdén et al., 2020; Sun et al., 2024), physical activity (Kaur et al., 2019; Nuzum et al., 2020; Erickson et al., 2022), mental health (Dogra et al., 2022) and social support (Sommerlad et al., 2019; Acoba, 2024).

Another factor that has recently gained prominence due to its significant influence on the cognitive performance of older adults is sleep quality. There is evidence that sleep quality is also affected by the aging process (Sakal et al., 2024). Older adults experience changes in sleep architecture, with a decrease in deep sleep and an increase in sleep fragmentation (Corral-Pérez et al., 2024; Li et al., 2016; Boyce et al., 2016; Wilckens et al., 2016; Guan et al., 2020). This results in difficulty falling asleep, frequent awakenings during the night, and a sense of unrefreshing sleep (Scullin and Bliwise, 2015; Wennberg et al., 2017). These sleep changes are intrinsically related to cognitive decline in older adults (Ourry et al., 2023; Wennberg et al., 2017).

Specifically, several studies have shown that poor sleep quality can contribute to a decline in cognitive functions such as memory, attention and processing speed (Guan et al., 2020), evidencing that poor sleep quality may predict worse cognitive performance (Kroeger & Vetrivelan, 2023). Some variables may modulate this relationship, as recently observed by Ourry et al. (2023), who found that cognitive reserve can modulate the relationship between sleep disturbances and memory performance in older adults, that is, day-to-day variability in sleep quality has also a significant impact on cognitive function in older adults. Several studies have found that greater inconsistency in sleep efficiency from day to day is associated with poorer cognitive performance (Guan et al., 2020). Other studies have found that variability in sleep efficiency is associated with an increased burden of β-amyloid, a biomarker of Alzheimer’s disease (Spira et al., 2014; Schwarz et al, 2021; Sun et al., 2024). Complementarily, Banerjee and Boro (2022) reported that poor sleep quality was associated with lower life satisfaction, both directly and indirectly, with mental health challenges mediating this relationship. All these findings suggest that consistency in sleep quality may be an important target for interventions aimed at preserving cognitive function and quality of life in older adults.

It is worth noting that this relationship between sleep quality and cognitive performance could be gender-dependent. In this regard, Curtis et al. (2024) showed that the relationship between sleep quality and spatial attentional orientation and processing speed was only obtained for women and not for men. Similarly, Wiranto et al. (2024) have shown that total sleep time is a predictor of executive functions and memory performance in older women but not in older men. Likewise, over the last decades it has been evidenced that women show a worse sleep quality (Sexton et al., 2017). Therefore, it is considered that the gender variable could be of interest in the present study with respect to the establishment of the relationship between the variables of cognitive performance, sleep quality and quality of life, variables that have not been studied in combination so far.

To date, sleep quality has been studied primarily as a variable associated with either the quality of life or cognitive performance of older adults, but these aspects have generally been examined independently. However, to our knowledge, none of these studies have explored the interaction between these two variables. Also, it has not been established whether sleep quality does not simply exert an associative but a moderating role between cognitive performance and the quality of life perceived by the individual, conditioning as well the impact that cognitive stimulation may have on healthy older adults. Taking this question into account, the primary aim of the present study is to evaluate how sleep quality influences the relationship between cognitive performance and quality of life on a sample of healthy older adults, as well as to evaluate differences between men and women in cognitive performance, sleep quality, and perceived quality of life. In order to address this aim, we propose the following hypotheses: (H1) Both sleep quality and cognitive performance will act as direct predictors of perceived quality of life, being positively associated with it; (H2) Sleep quality will act as a moderator variable between cognitive performance and perceived quality of life, more specifically, the poorer sleep quality, the higher importance of cognitive performance for a better quality of life; (H3) Taking into account that women suffer of poorer sleep quality, women will evidence a greater moderation role of sleep quality on the relation between cognitive performance and perceived quality of life than men.

## Methods

### Participants

The study initially included 57 participants, selected through convenience sampling from a university program for older adults in a Spanish university, which is aimed at individuals aged 55 and above. During the experimental procedure, 7 participants opted to withdraw, resulting in a final sample of 50 participants (mean age = 65.48 ± 5.68 years; 38 women, 74.5%). Eligibility criteria required participants to be over 55 years old, free from sensory or motor impairments that could interfere with completing the questionnaires. The study adhered to the ethical principles of the Declaration of Helsinki and received approval from the Ethics Committee of the University of Valencia (Reference: 2023-PSILOG-2558999).

### Instruments

#### Cognitive performance

The Fototest (Carnero-Pardo et al., 2007) is a brief cognitive screening tool designed to evaluate memory, language, and visual recognition abilities. It consists of three subtests: naming, free recall, and category-cued recall, all based on five photographs of common objects. Scores range from 0 to 50, with higher scores indicating better cognitive performance. The Fototest has demonstrated strong validity and reliability in assessing cognitive functioning in older adults (Carnero-Pardo et al., 2007).

#### Sleep quality

The Pittsburgh Sleep Quality Index (PSQI) (Buysse et al., 1989) is a self-report questionnaire assessing sleep quality and disturbances over the past month. It consists of 19 items grouped into seven components: subjective sleep quality, sleep latency, sleep duration, habitual sleep efficiency, sleep disturbances, use of sleep medication, and daytime dysfunction. These components generate a global score ranging from 0 to 21, with higher scores indicating poorer sleep quality. A global score above 5 suggests significant sleep disturbances. The PSQI has been extensively validated and is widely used in clinical and research settings to assess sleep health.

#### Quality of life

The Spanish version (Badia et al., 1999) of European Quality of Life 5 Dimensions (EuroQoL-5D) (Balestroni & Bertolotti, 2015) is a self-report questionnaire used to assess the perceived quality of life. In this study, quality of life has been evaluated using the indicator of perceived general health, the Visual Analogue Scale of the EuroQol-5D Questionnaire, which is a single item to which the participants must answer with a number in units ranging between the minimum value of 0 (representing “the worst imaginable state of health”) and the maximum value of 100 (representing “the best imaginable state of health”).

### Procedure

The study sessions were conducted from Monday to Thursday between 9:00 AM and 12:45 PM, with each session lasting approximately 45 minutes to 1 hour. Participants were contacted and scheduled to meet at a designated location within the educational center where the study was conducted. Upon arrival, participants were accompanied to the laboratory, where the general procedure was explained, and informed consent forms were read and signed. Subsequently, participants completed a battery of questionnaires, including the Fototest, the PSQI, and the EuroQol-5D Questionnaire.

### Statistical analyses

Outliers were identified using the 2.5 standard deviation method. Normality was assessed using the Kolmogorov-Smirnov test with Lilliefors correction. Descriptive statistics were calculated for the entire sample and stratified by gender, with ANOVA tests performed to evaluate differences between men and women in cognitive performance (Fototest), sleep quality (PSQI), and perceived quality of life (EuroQol-5D). Correlation analyses were conducted to examine relationships between cognitive performance, sleep quality, and quality of life across the total sample and separately by gender. Pearson’s correlation coefficients were used to determine the strength and direction of these relationships. Multiple linear regression analyses were conducted to assess whether cognitive performance and sleep quality predicted quality of life, controlling for gender as a covariate. A moderation analysis was performed to evaluate whether sleep quality moderated the relationship between cognitive performance and quality of life. The Johnson-Neyman technique was applied to identify regions of significance, exploring conditional effects at different levels of sleep quality. The significance level (α) was set at .05, and partial eta squared (η^2^p) was used to indicate effect sizes where applicable. All analyses were performed using IBM SPSS Statistics 25.

## Results

### Descriptive Statistics

The general descriptive statistics for the sample are presented in Table 1. These variables were also compared by gender to examine potential differences between men and women. As shown in Table 1, there were significant gender differences in cognitive performance and sleep quality evidenced by the scores of Fototest and PSQI, respectively. Compared to women, men scored lower on the cognitive test and reported better sleep quality. No significant differences were observed in quality of life as shown by EuroQoL-5D score. However, the disproportionate gender distribution in the sample (71.43% women and 28.57% men) requires caution when interpreting these results, as this imbalance could increase the likelihood of a Type II error.

**Table 1.**
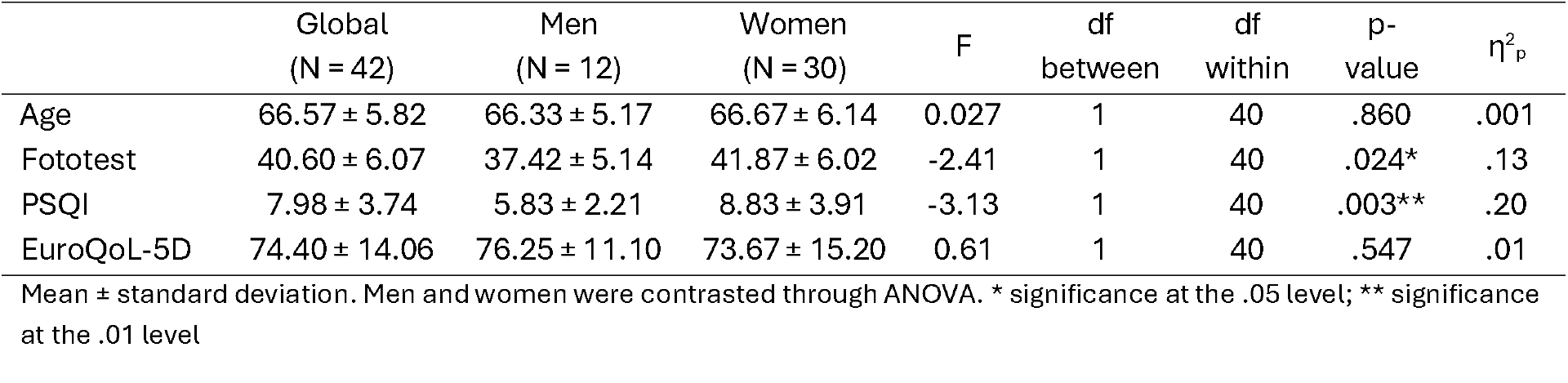
Descriptive statistics of the scores obtained from the Fototest, PSQI, and EuroQoL-5D instruments, & comparison between gender.

| | Global<br>(N = 42) | Men<br>(N = 12) | Women<br>(N = 30) | F | df<br>between | df<br>within | p-<br>value | $\eta^2_p$ |
| --- | --- | --- | --- | --- | --- | --- | --- | --- |
| Age | 66.57 ± 5.82 | 66.33 ± 5.17 | 66.67 ± 6.14 | 0.027 | 1 | 40 | .860 | .001 |
| Fototest | 40.60 ± 6.07 | 37.42 ± 5.14 | 41.87 ± 6.02 | -2.41 | 1 | 40 | .024* | .13 |
| PSQI | 7.98 ± 3.74 | 5.83 ± 2.21 | 8.83 ± 3.91 | -3.13 | 1 | 40 | .003** | .20 |
| EuroQoL-5D | 74.40 ± 14.06 | 76.25 ± 11.10 | 73.67 ± 15.20 | 0.61 | 1 | 40 | .547 | .01 |

Mean ± standard deviation. Men and women were contrasted through ANOVA. \* significance at the .05 level; \*\* significance at the .01 level

### Interrelation between Cognitive performance, Sleep Quality & Quality of life

A correlation analysis was performed for cognitive performance, sleep quality and quality of life, variables obtained from the Fototest, PSQI, and, EuroQoL-5D instruments, respectively. As can be seen, a medium positive correlation was found between cognitive performance and perceived quality of life, while a negative medium correlation was observed between quality of life and sleep quality; that is, the higher the quality of life, the less sleep problems are reported, since the interpretation of the PSQI scale is inverse(the values of Pearson’s correlation coefficient and p-values are detailed in the Heatmap in Figure 1). The relationship between cognitive performance and sleep quality was weaker and not significant.

**Figure 1.**
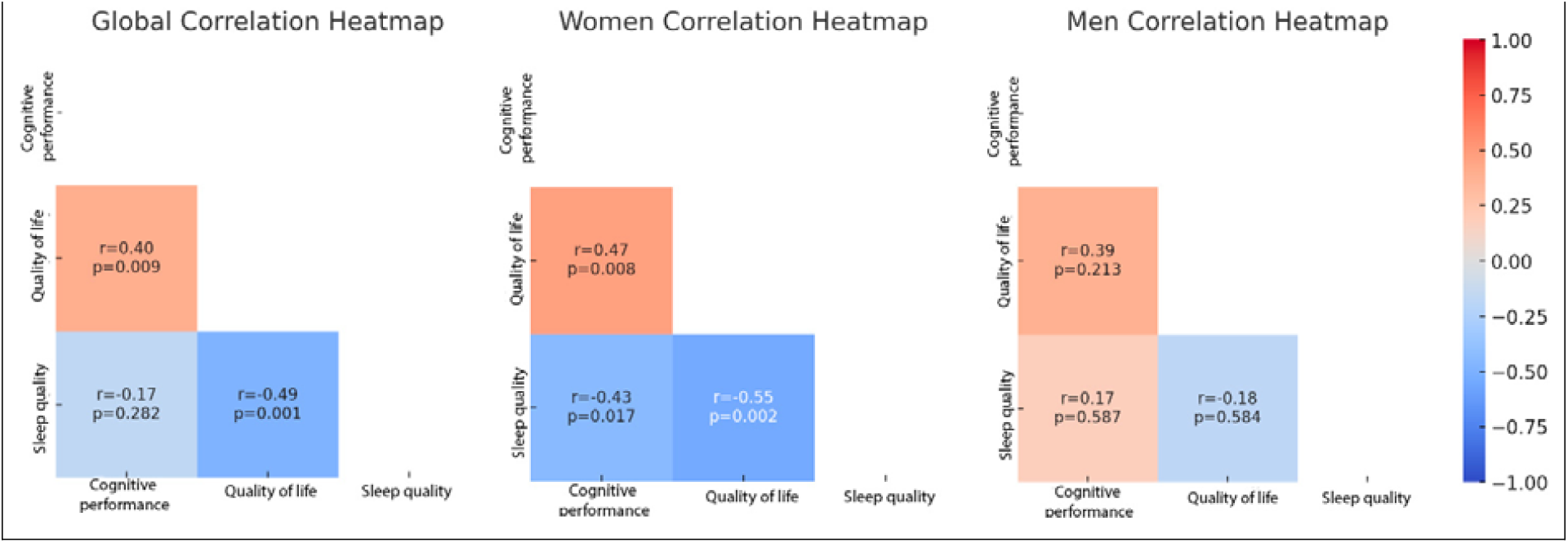
Heat map of the correlations between cognitive performance, sleep quality & quality of life, global and by gender

These correlations were also separately tested for each gender. As also seen in Figure 1, women maintained a similar pattern of relationships between the main variables. Once again, a medium positive correlation was found between cognitive performance and perceived quality of life, and a negative correlation was observed between quality of life and sleep quality. However, in this case, a significant and negative relationship was also found between cognitive performance and sleep quality in women. For men, none of the correlations reached statistical significance. Although there was a trend suggesting a potential positive correlation between cognitive performance and quality of life, and a negative trend between quality of life and sleep quality, these did not meet the threshold for significance (*p* > 0.05).

### Regression analyses

To assess whether cognitive performance and sleep quality could predict perceived quality of life, controlling for the potential influence of gender, a multiple linear regression analysis was performed. The predictors included in the model were the total scores from the Fototest and PSQI instruments, with gender as a covariate. Quality of life was the dependent variable.

The overall regression model was significant, *F*(3, 38) = 6.64, *p* =.001, explaining 34.4% of the variance in quality of life (R^2^ = 0.344), with an adjusted R^2^ = 0.29. Cognitive performance was a significant positive predictor of quality of life (*B* = 0.80, *SE* = 0.34, *t* = 2.34, *p* = .025), indicating that higher scores on the Fototest were associated with better perceived quality of life. Sleep quality was a significant negative predictor (*B* = −1.55, *SE* = 0.56, *t* = −2.75, *p* = .009), showing that poorer sleep quality was linked to lower perceived quality of life. However, the inclusion of gender as a covariate did not yield a significant effect (*B* = 1.51, *SE* = 4.82, *t* = 0.31, *p* = .756), suggesting that gender did not significantly influence the relationship between cognitive performance, sleep quality, and perceived quality of life in this sample. In conclusion, both cognitive performance and sleep quality significantly predicted perceived quality of life, while gender did not play a significant role in moderating these effects.

### Moderation analysis

A moderation analysis was conducted to examine whether sleep quality moderates the relationship between cognitive performance and perceived quality of life, again controlling gender as a covariate.

The overall model was significant, explaining approximately 42.2% of the variance in quality of life (R2 = 0.42), *F*(4, 37) = 6.76, *p* < .001. As seen in the previous section, gender did not yield a significant effect (*B* = -1.90, *SE* = 4.82, *t* = -0.39, *p* = .93), and sleep quality remained a significant negative predictor of quality of life (*B* = −9.99, *SE* = 3.81, *t* = −2.63, *p* = .012). However, unlike in the regression analysis where cognitive performance had a significant direct effect on quality of life, in this model the direct effect of cognitive performance was no longer significant (*B* = −0.93, *SE* = 0.84, *t* = −1.11, *p* = .276). This suggests that, when the interaction between cognitive performance and sleep quality is included, cognitive performance alone is no longer sufficient to predict quality of life without considering the moderating role of sleep quality. The interaction between cognitive performance and sleep quality was found to be significant (*B* = 0.21, *SE* = 0.09, *t* = 2.24, *p* = .031), indicating that the relationship between cognitive performance and quality of life is moderated by sleep quality. This interaction accounted for an additional 7.84% of the variance in quality of life (R^2^change = 0.078), *F*(1, 37) = 5.02, *p* =.031, which was not captured in the previous regression analysis.

To further explore this moderation effect, the Johnson-Neyman technique was applied to identify regions of significance for the interaction. The results showed that the effect of cognitive performance on quality of life becomes significant when PSQI scores are above 7.54. As illustrated in Figure 2, the positive relationship between cognitive performance and quality of life is significant when sleep quality is moderate to poor (PSQI > 7.54). For individuals with higher PSQI scores (indicating poorer sleep quality), better cognitive performance is associated with higher quality of life. Conversely, when PSQI scores are lower (indicating better sleep quality), cognitive performance does not significantly affect quality of life.

**Figure 1.**
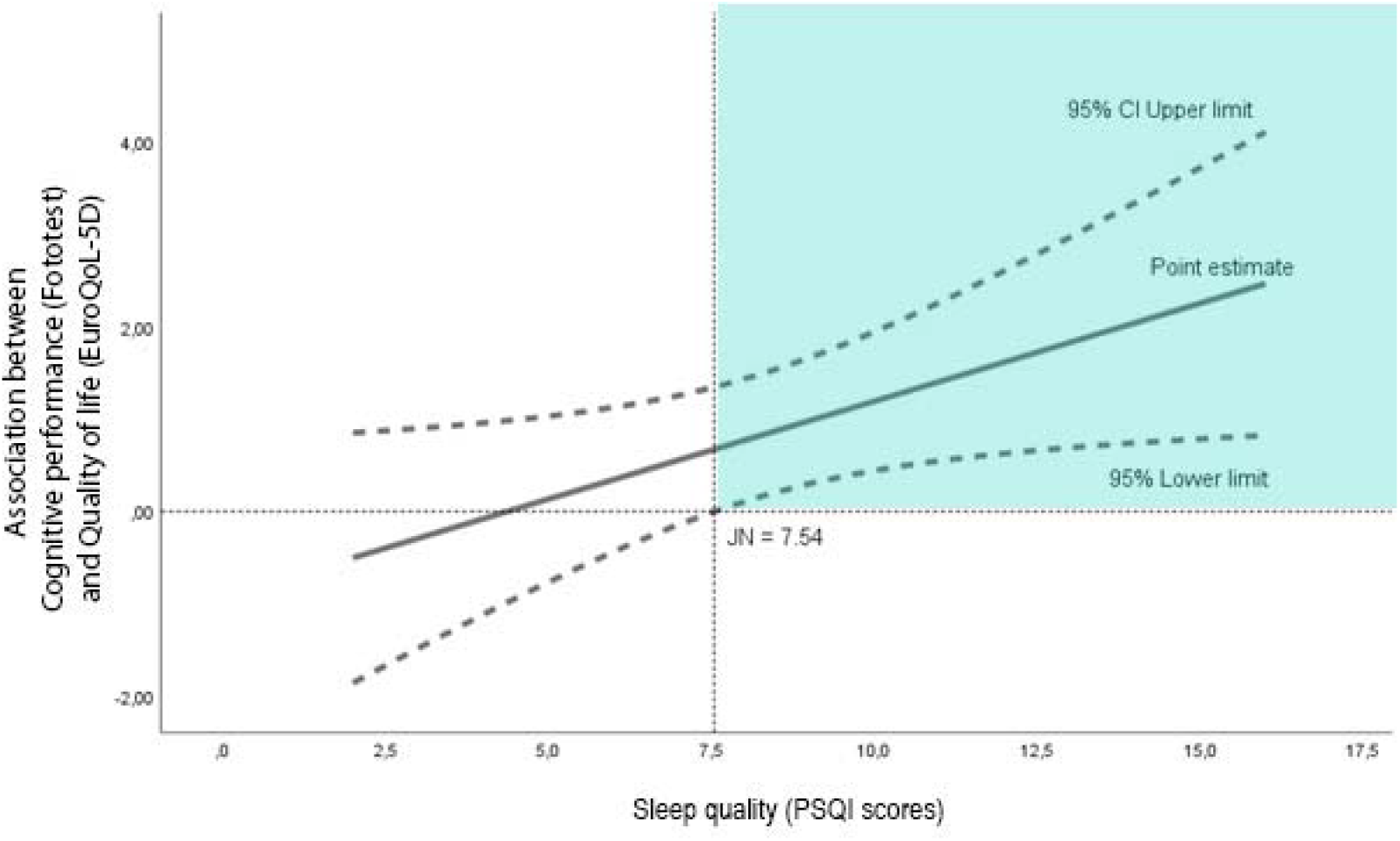
Johnson-Neyman graph. Graph of the conditional association between cognitive performance and quality of life, as a linear function of sleep quality including the Johnson-Neyman transition point (JN). The JN point is where the confidence interval around the condition effect interests zero on the y-axis. Thus, the shaded quadrant is the region of significance, i.e. those values of sleep quality for which the association between cognitive performance and quality of life was significant.

In summary, these findings suggest that sleep quality not only directly influences perceived quality of life but also moderates the relationship between cognitive performance and quality of life. While cognitive performance was previously a direct predictor of quality of life, its effect seems to depend on an individual’s level of sleep quality. The Johnson-Neyman plot (Figure 2) illustrates this conditional effect across different levels of sleep quality.

## Discussion

The primary aim of this study was to assess whether sleep quality plays a moderating role, rather than merely serving as an associative or predictive factor, in the relationship between cognitive performance and quality of life in healthy older adults. The results confirmed our hypotheses. On one hand, cognitive performance and sleep quality acted as predictors of perceived quality of life, both directly influencing the level of quality of life. However, even though cognitive performance was a direct predictor of quality of life, this effect depended on the sleep quality experienced by the individuals. These results will be further addressed below.

Primarily, and as expected, correlations analyses showed an association between participants’ cognitive performance, sleep quality and perceived quality of life. More specifically, perceived quality of life was positively related to participants’ cognitive performance, and negatively related to their reported sleep problems. These results are consistent with previous literature since, as shown by Meng et al. (2023), cognitive function in adulthood has a crucial impact on their quality of life being one of the influential factors. In this line, participants with cognitive impairment or diminished cognitive performance show a decreased level of perceived quality of life (Hartmann et al., 2021). Even more, higher cognitive reserve maintained an optimal cognitive performance playing a moderating role in how poor sleep quality affects cognitive function in older population and, thus, moderating the impact on wellbeing (Ourry et al., 2023). On the other hand, although in the overall results we did not obtain significant data regarding the relationship between cognitive performance and sleep quality, this relationship was corroborated when the analysis was performed only on women. These results would go in the direction of what was found by Dzierzewski et al. (2022), who proved the link between sleep quality and cognitive function, and stated that poor sleep quality is related to cognitive decline. Even more, when using the Mini-Mental State Examination (MMSE) as a cognitive evaluation, it decreased with increased PSQI score, that is, the worse the sleep quality, the worse the cognitive function (Joo et al., 2021; Meng et al., 2023; Künstler et al., 2023).

With regard to the predictive power of cognitive performance and sleep quality variables on participants’ perceived quality of life, our hypothesis is confirmed, both showing a direct effect in the regression model tested on quality of life. In this context, our results support previous findings such as those of Meng et al. (2023), who showed that cognitive performance is an important predictor of quality of life in the elderly. Likewise, several studies have also corroborated the predictive value of sleep quality on quality of life (Gothe et al., 2020; Leng et al., 2020). In addition, Guan et al. (2020) found that there exists a relation between sleep disturbance and cognitive processing; specifically, there exists an association between sleep continuity and processing speed in older adults aged over 65 years which was previously evidenced by Blackwell et al. (2006). Long-term sleep disturbances can impair functioning across multiple life domains, potentially requiring more sustained efforts to improve sleep hygiene (Mukherjee et al., 2024). Moreover, Guan et al. (2020) detailed that in late life, the estrogen level reduces and fluctuates more in women, while the estrogen level is relatively stable via the metabolism of testosterone in men. Therefore, although using gender as a covariate, our analyses have shown that its weight in the prediction model was not relevant, this could be due to the small number of men in the sample of participants in the present study.

Furthermore, and after executing a moderation analysis, the results have pointed to interesting conclusions indicating that, in fact, cognitive performance influences quality of life depending on the quality of sleep experienced by the participant. In other words, when considering the interaction of cognitive performance and sleep quality, it is evident that cognitive performance alone no longer possesses sufficient predictive value with respect to perceived quality of life if the moderating role of sleep quality is not taken into account. Therefore, in the present study, a relevant result for literature has been evidenced and suggests that the relationship between cognitive performance and quality of life is influenced by sleep quality. Specifically, the positive relationship between cognitive performance and quality of life is significant when sleep quality is moderate to poor, whereas when PSQI scores are lower (indicating better sleep quality), cognitive performance does not significantly affect quality of life. In this context, Meng et al. (2023) established a relation between cognition and sleep quality, but our results contribute with a cut-off point that can be useful for interpreting this relation and the moderating role of sleep quality. Therefore, these results provide a basis on which to build *ad hoc* cognitive intervention programs, which are known to be beneficial (Krivanek et al., 2021), considering the sleep quality variable as a moderator. That is, despite emphasizing in previous studies the weight of the sleep quality variable as a mediating variable on cognitive function and quality of life (Guan et al., 2020; Guo et al., 2022; Liu et al., 2022; Sun et al., 2024), it had never been considered a moderation model such as the one proposed in the present study.

Moreover, the results obtained about the gender variable seem to point in the hypothesized direction, observing that the sample of women in the present study showed worse sleep quality and worse quality of life than the sample of men. Specifically, women presented a positive relationship between cognitive performance and perceived quality of life, while a negative correlation was obtained between quality of life and quality of sleep. A negative correlation was also obtained between cognitive performance and sleep quality. In contrast, men did not show significant results in any of the correlations tested.

In general, these results are in agreement with previous data that has been studied independently indicating that women tend to show more evident sleep problems such as total sleep time, sleep latency, wake after sleep onset, sleep efficiency (Yaffe et al., 2007; Diem et al., 2016) and some sleep architecture disruption on slow-wave sleep and rapid eye movement sleep (Blackwell et al., 2006), which in turn may interfere with their cognitive performance and quality of life (Guan et al., 2020; Casagrande et al., 2022). In contrast of the present study, all these results studied the variables by pairs or separately. Regarding the effects observed in older women in our study, these are in line with those recently obtained by Wiranto et al. (2024). In their study, they showed that poorer self-reported sleep quality was associated with poorer cognitive performance in older women, but no such effect was observed in older men, where higher cognitive performance was obtained with more severe self-reported sleep quality.

Some of the limitations of the present study are, the small number of men in the sample of the study, so the conclusions drawn from these results should be taken with caution. To this end, it would be advisable to use larger samples with an equal number of men and women in the future in order to be able to contrast possible differences between the two groups. This aspect could also entail contrasting whether the sleep quality factor is relevant in both men and women with respect to cognitive performance and its influence on quality of life. In this sense, and unlike what is evidenced in the literature, it would be recommendable to use the sleep quality variable as a moderating factor in future studies since, according to our results, this variable explains a relevant percentage of the variance of the model between cognitive performance and quality of life perceived by healthy older adults.

Despite these limitations, this study has provided valuable evidence on how the variables of cognitive performance, sleep quality and perceived quality of life are related to each other in healthy older adults. Specifically, sleep quality not only directly influences quality of life, which was already intuited in previous studies, but also moderates the relationship between cognitive performance and quality of life. Thus, while cognitive performance was initially a direct predictor of quality of life, according to the present results, it seems that this relationship depends on the quality of sleep of each individual. Consequently, it would be interesting to promote further research on the significance of this variable when implementing a cognitive intervention program in a sample of older adults, as well as to have a forecast of the impact that such intervention will have on the sample considering the sleep quality of the individuals.

## Data Availability

All data produced in the present study are available upon reasonable request to the authors

## Acknowledgements

This work was supported by the grant PID2022-138021OA-I00 funded by MCIN/AEI/10.13039/501100011033/ and, by “ERDF A way of making Europe”.

## References

Acoba, E. F. (2024). Social support and mental health: the mediating role of perceived stress. Frontiers in Psychology, 15. 10.3389/FPSYG.2024.1330720

Badia, X., Roset, M., Montserrat, S., Herdman, M., & Segura, A. (1999). The Spanish version of EuroQol: a description and its applications. European Quality of Life scal. Medicina Clínica; 112 Suppl1:79–85.

Băjenaru, L., Balog, A., Dobre, C., Drăghici, R., & Prada, G.I. (2022). Latent profile analysis for quality of life in older patients. BMC Geriatr. 22(1):848. 10.1186/s12877-022-03518-1

Balestroni, G., & Bertolotti, G. (2015). “EuroQol-5D (EQ-5D): An Instrument for Measuring Quality of Life”. Monaldi Archives for Chest Disease 78(3). 10.4081/monaldi.2012.121

Banerjee, S., & Boro, B. Analysing the role of sleep quality, functional limitation and depressive symptoms in determining life satisfaction among the older Population in India: a moderated mediation approach. BMC Public Health 22, 1933 (2022). 10.1186/s12889-022-14329-9

Blackwell, T., Yaffe, K., Ancoli-Israel, S., Schneider, J.L., Cauley, J.A., Hillier, T.A., Fink, H.A., & Stone, K.L. (2006). Poor sleep is associated with impaired cognitive function in older women: The study of osteoporotic fractures. J Gerontol A Biol Sci Med Sci 61(4):405–10. 10.1093/gerona/61.4.405

Boyce, R., Glasgow, S.D., Williams, S., & Adamantidis, A. (2016). Causal evidence for the role of REM sleep theta rhythm in contextual memory consolidation. Science 352(6287):812–6. 10.1126/science.aad5252

Brandt, L., Liu, S., Heim, C., & Heinz, A. (2022). The effects of social isolation stress and discrimination on mental health. Transl Psychiatry 12(1):398. 10.1038/s41398-022-02178-4

Brett, C.E., Dykiert, D., Starr, J.M., & Deary, I.J. (2019). Predicting change in quality of life from age 79 to 90 in the Lothian Birth Cohort 1921. Qual Life Res. 28(3):737–49.

Buysse, D.J., Reynolds, C.F., Monk, T.H., Berman, S.R., & Kupfer, D.J. (1989). The Pittsburgh sleep quality index: A new instrument for psychiatric practice and research. Psychiatry Res. 28(2):193–213. 10.1016/0165-1781(89)90047-4

Campos, A.C., Ferreira e Ferreira, E., Vargas, A.M., & Albala, C. (2016). Aging. Gender and quality of life (AGEQOL) study: factors associated with good quality of life in older Brazilian community-dwelling adults. Health Qual Life Outcomes. 12:166. 10.1186/s12955-014-0166-4

Carnero Pardo, C., Sáez-Zea, C., Montiel Navarro, L., Del Saz, P., Feria Vilar, I., Pérez Navarro, M.J., et al. (2007). Utilidad diagnostica del Test de las Fotos (Fototest) en deterioro cognitivo y demencia. Neurologia. 22(10):860–9.

Casagrande, M., Forte, G., Favieri, F., & Corbo, I. (2022). Sleep Quality and Aging: A Systematic Review on Healthy Older People, Mild Cognitive Impairment and Alzheimer’s Disease. Int. J. Environ. Res. Public Health, 19(14):8457. 10.3390/ijerph19148457

Christopher, G., & Facal, D. (2023). Editorial: Emotion regulation and mental health in older adults. Frontiers in Psychology, 14, 1173314. 10.3389/FPSYG.2023.1173314/BIBTEX

Corral-Pérez, J., Vázquez-Sánchez, M.Á., Casals-Sánchez, J.L., Contreras-García, F.J., Costilla, M. & Casals, C. (2024). A 6-month educational program improves sleep behaviour in community-dwelling frail older adults: A randomised controlled trial. Sleep Med. 121:196–202. 10.1016/j.sleep.2024.07.011

Curtis, A.F., Costa, A.N., Musich, M., Schmiedeler, A., Jagannathan, S., Connell, M., Atkinson, A., Miller, M.B., & McCrae, C.S. (2024). Sex as a moderator of the sleep and cognition relationship in middle-aged and older adults: A preliminary investigation. Behav Sleep Med 22(1):14–27. 10.1080/15402002.2023.2177293

Davis, J.C., Bryan, S., Best, J.R., Li, L.C., Hsu, C.L., Gomez, C., et al. (2015). Mobility predicts change in older adults’ health-related quality of life: evidence from a Vancouver falls prevention prospective cohort study. Health Qual Life Outcomes. 13:101. 10.1186/s12955-015-0299-0

Diem, S.J., Blackwell, T.L., Stone, K.L., Yaffe, K., Tranah, G., Cauley, J.A., & Ensrud, K.E. (2016). Measures of sleep–wake patterns and risk of mild cognitive impairment or dementia in older women. Am J. Geriatr. 24(3):248–58. 10.1016/j.jagp.2015.12.002

Dogra, S., Dunstan, D.W., Sugiyama, T., Stathi, A., Gardiner, P.A., & Owen, N. (2022). Active Aging and Public Health: Evidence, Implications, and Opportunities. Annu. Rev. Public Health, 43, 439–459. 10.1146/annurev-publhealth-052620-091107

Dzierzewski, J.M., Perez, E., Ravyts, S.G., & Dautovich, N. (2022). Sleep and Cognition: A narrative review focused on older adults. Sleep Med Clin. 17(2):205–222. 10.1016/j.jsmc.2022.02.001

Erickson, K.I., Donofry, S.D., Sewell, K.R., Brown, B.M., & Stillman, C.M. (2022). Cognitive Aging and the Promise of Physical Activity. Annu Rev Clin Psychol. 18:417–442. 10.1146/annurev-clinpsy-072720-014213

Gothe, N.P., Ehlers, D.K., Salerno, E.A., Fanning, J., Kramer, A.F., & McAuley, E. (2020). Physical activity, sleep and quality of life in older adults: Influence of physical, mental and social well-being. Behav. Sleep Med. 18(6):797–808. 10.1080/15402002.2019.1690493

Guan, Q., Hu, X., Ma, N., He, H., Duan, F., Li, X., Luo, Y., & Zhang, H. (2020). Sleep Quality, Depression, and Cognitive Function in Non-Demented Older Adults. J Alzheimers Dis. 76(4):1637–1650. 10.3233/JAD-190990

Guo, H., Zhangm Y., Wang, Z., & Shen, H. (2022). Sleep Quality Partially Mediate the Relationship Between Depressive Symptoms and Cognitive Function in Older Chinese: A Longitudinal Study Across 10 Years. Psychol Res Behav Manag. 15:785–799. 10.2147/PRBM.S353987

Hartmann, J., Roßmeier, C., Riedl, L., Dorn, B., Fischer, J., Slawik, T., et al. (2021). Quality of life in Advanced Dementia with Late Onset, Young Onset, and very young onset. J Alzheimers Dis., 80(1):283–97. 10.3233/JAD-201302

He, C., Kong, X., Li, J., Wang, X., Chen, X., Wang, Y., Zhao, Q., & Tao, Q. (2023). Predictors for quality of life in older adults: network analysis on cognitive and neuropsychiatric symptoms. BMC Geriatr. 23(1):850. 10.1186/s12877-023-04462-4

Joo, H.J., Joo, J.H., Kwon, J. et al. (2021). Association between quality and duration of sleep and subjective cognitive decline: a cross-sectional study in South Korea. Sci Rep 11, 16989 10.1038/s41598-021-96453-x

Kaur, S., Banerjee, N., Miranda, M., Slugh, M., Sun-Suslow, N., McInerney, K.F., Sun, X., Ramos, A.R., Rundek, T., Sacco, R.L., & Levin, B.E. (2019). Sleep quality mediates the relationship between frailty and cognitive dysfunction in non-demented middle aged to older adults. Int Psychogeriatr. 31(6):779–788. 10.1017/S1041610219000292

Krivanek, T. J., Gale, S.A., McFeeley, B.M., Nicastri, C.M., & Daffner, K.R. (2021). Promoting Successful Cognitive Aging: A Ten-Year Update. Journal of Alzheimer’s Disease, 81(3), 871–920. 10.3233/JAD-201462

Kroeger, D., & Vetrivelan, R. (2023). To sleep or not to sleep - Effects on memory in normal aging and disease. Aging Brain. 3:100068. 10.1016/j.nbas.2023.100068

Künstler, E.C.S., Bublak, P., Finke, K., Koranyi, N., Meinhard, M., Schwab, M., & Rupprecht, S. (2023). The Relationship Between Cognitive Impairments and Sleep Quality Measures in Persistent Insomnia Disorder. Nat Sci Sleep. 15:491–498 10.2147/NSS.S399644

Lapid, M.I., Rummans, T.A., Boeve, B.F., McCormick, J.K., Pankratz, V.S., Cha, R.H., Smith, G.E., Ivnik, R.J., Tangalos, E.G., & Petersen, R.C. (2011). What is the quality of life in the oldest old? Int Psychogeriatr. 23(6):1003–10. 10.3233/JAD-201462

Leng, M., Yin, H., Zhang, P., Jia, Y., Hu, M., Li, G., Wang, C., & Chen, L. (2020). Sleep Quality and Health-Related Quality of Life in Older People With Subjective Cognitive Decline, Mild Cognitive Impairment, and Alzheimer Disease. J Nerv Ment Dis. 208(5):387–396. 10.1097/NMD.0000000000001137

Li, Y., Liu, H., Weed, J.G., Ren, R., Sun, Y., Tan, L., & Tang, X. (2016). Deficits in attention performance are associated with insufficiency of slow-wave sleep in insomnia. Sleep Med. 24:124–130. 10.1016/j.sleep.2016.07.017 Erratum in: Sleep Med. 84:425. 10.1016/j.sleep.2021.04.008 Erratum in: Sleep Med. 84:424. 10.1016/j.sleep.2021.05.018

Liu, X., Xia, X., Hu, F., Hao, Q., Hou, L., Sun, X., Zhang, G., Yue, J., & Dong, B. (2022). The mediation role of sleep quality in the relationship between cognitive decline and depression. BMC Geriatr. 22(1):178. 10.1186/s12877-022-02855-5

Lövdén, M., Fratiglioni, L., Glymour, M.M., Lindenberger, U., & Tucker-Drob, E.M. (2020). Education and cognitive functioning across the life span. Psychol Sci Public Interest. 21(1):6–41. 10.1177/1529100620920576

Meng, W., Gao, T., Zhong, Y., & Ge, L. (2023). Association Between Sleep and Cognition of Older Adults in Rural Areas: A Cross-Sectional Study. Inquiry., 60:469580231171820. 10.1177/00469580231171820

Mukherjee, U., Sehar, U., Brownell, M., & Reddy, P.H. (2024). Mechanisms, consequences and role of interventions for sleep deprivation: Focus on mild cognitive impairment and Alzheimer’s disease in elderly. Ageing Res Rev. 100:102457. 10.1016/j.arr.2024.102457

Nuzum, H., Stickel, A., Corona, M., Zeller, M., Melrose, R.J., & Wilkins, S.S. (2020). Potential Benefits of Physical Activity in MCI and Dementia. Behav Neurol. 12:2020:7807856. 10.1155/2020/7807856

Ourry, V., Rehel, S., André, C., Mary, A., Paly, L., Delarue, M., Requier, F., Hendy, A., Collette, F., Marchant, N.L., Felisatti, F., Palix, C., Vivien, D., de la Sayette, V., Chételat, G., Gonneaud, J., & Rauchs, G. (2023). Medit-Ageing Research Group. Effect of cognitive reserve on the association between slow wave sleep and cognition in community-dwelling older adults. Aging (Albany NY).15(18):9275–9292. 10.18632/aging.204943

Rainey-Smith, S.R., Mazzucchelli, G.N., Villemagne, V.L., Brown, B.M., Porter, T., Weinborn, M., & Laws, S.M. (2018). Genetic variation in Aquaporin-4 moderates the relationship between sleep and brain A-amyloid burden. Trans. Psychiatry 8(1):47. 10.1038/s41398-018-0094-x

Sakal, C., Li, T., Li, J., Yang, C., & Li, X. (2024). Association Between Sleep Efficiency Variability and Cognition Among Older Adults: Cross-Sectional Accelerometer Study. JMIR Aging. 7:e54353. 10.2196/54353

Schwarz, C., Lange, C., Benson, G.S., Horn, N., Wurdack, K., Lukas, M., Buchert, R., Wirth, M., & Flöel, A. (2021). Severity of Subjective Cognitive Complaints and Worries in Older Adults Are Associated with Cerebral Amyloid-β Load. Front Aging Neurosci.;13:675583. 10.3389/fnagi.2021.675583

Scullin, M.K., & Bliwise, D.L. (2015). Sleep, cognition, and normal aging: integrating a half century of multidisciplinary research. Perspect Psychol Sci. 10(1):97–137. 10.1177/1745691614556680

Sexton, C.E., Zsoldos, E., Filippini, N., Griffanti, L., Winkler, A., Mahmood, A., Allan, C.L., Topiwala, A., Kyle, S.D., Spiegelhalder, K., Singh-Manoux, A., Kivimaki, M., Mackay, C.E., Johansen-Berg, H., & Ebmeier, K.P. (2017). Associations between self-reported sleep quality and white matter in community-dwelling older adults: A prospective cohort study. Hum Brain Mapp. 38(11):5465–5473. 10.1002/hbm.23739

Shrestha, S., Stanley, M.A., Wilson, N.L., Cully, J.A., Kunik, M.E., Novy, D.M., Rhoades, H.M., & Amspoker, A.B. (2015). Predictors of change in quality of life in older adults with generalized anxiety disorder. Int Psychogeriatr. 27(7):1207–15. 10.1017/S1041610214002567

Sommerlad, A., Sabia, S., Singh-Manoux, A., Lewis, G., & Livingston, G. (2019). Association of social contact with dementia and cognition: 28-year follow-up of the Whitehall II cohort study. PLoS Med. 16(8):e1002862. 10.1371/journal.pmed.1002862

Spira, A.P., Yager, C., Brandt, J., Smith, G.S., Zhou, Y., Mathur, A., Kumar, A., Brašić, J.R., Wong, D.F., & Wu. M.N. (2014). Objectively Measured Sleep and β-amyloid Burden in Older Adults: A Pilot Study. SAGE Open Med.; 2:2050312114546520. 10.1177/2050312114546520

Stone, A.A., Schwartz, J.E., Broderick, J.E., & Deaton, A. (2010). A snapshot of the age distribution of psychological well-being in the United States. Proc Natl Acad Sci U S A. 107(22):9985–90. 10.1073/pnas.1003744107

Sun, M., Zhang, Q., Han, Y., & Liu, J. (2024). Sleep Quality and Subjective Cognitive Decline among Older Adults: The Mediating Role of Anxiety/Depression and Worries. J Aging Res.; 2024:4946303. 10.1155/2024/4946303

Wennberg, A.M.V., Wu, M.N., Rosenberg, P.B., & Spira, A.P. (2017). Sleep Disturbance, Cognitive Decline, and Dementia: A Review. Semin Neurol. 37(4):395–406. 10.1055/s-0037-1604351

Wilckens, K.A., Hall, M.H., Nebes, R.D., Monk, T.H., & Buysse, D.J. (2016). Changes in cognitive performance are associated with changes in sleep in older adults with insomnia. Behav Sleep Med 14(3):295–310. 10.1080/15402002.2014.1002034

Wiranto, Y., Siengsukon, C., Mazzotti, D.R., Burns, J.M., & Watts, A. (2024). Sex differences in the role of sleep on cognition in older adults. Sleep Adv, 5(1):zpae066. 10.1093/sleepadvances/zpae066

Yaffey, K., Blackwell, T., Barnes, D.E., Ancoli-Israel, S., & Stone, K.L. (2007). Preclinical cognitive decline and subsequent sleep disturbance in older women. Neurology. 69(3):237–42. 10.1212/01.wnl.0000265814.69163.da

